# Assessment of Sleep Hygiene Practices and Its Effects on Sleep Quality Among Medical Students at UMST, Sudan

**DOI:** 10.64898/2026.04.26.26351757

**Authors:** Mohammed Atif Abdelmajeed, Babiker Mohamed Ali Rahamtalla

**Affiliations:** Faculty of Medicine, University of Medical Sciences and Technology (UMST), Khartoum, Sudan; Head of Department of Community Medicine, UMST, Khartoum, Sudan

**Keywords:** sleep hygiene, sleep quality, medical students, Sudan, PSQI, SHI, cross-sectional study

## Abstract

**Background:** Medical students face demanding academic schedules and elevated stress levels, predisposing them to poor sleep quality. Sleep hygiene — a set of behavioural and environmental practices aimed at optimising sleep — has been identified as a modifiable determinant of sleep quality, yet its role among medical students in Sudan remains unstudied.

**Objectives:** To assess current sleep hygiene practices among medical students at UMST and determine their association with sleep quality outcomes.

**Methods:** A facility-based cross-sectional study was conducted at UMST among 240 medical students from three academic batches (3rd, 4th, and 5th year), selected via stratified random sampling. Data were collected using two validated self-administered instruments: the Pittsburgh Sleep Quality Index (PSQI) and the Sleep Hygiene Index (SHI). Descriptive statistics, independent sample t-tests, one-way ANOVA, chi-square tests, Pearson correlation, and binary logistic regression were performed using SPSS version 23.

**Results:** Poor sleep quality (PSQI >5) was prevalent in 72.1% of participants (mean PSQI 7.25 ± 2.66), and poor sleep hygiene (SHI >16) in 92.5% (mean SHI 27.1 ± 7.9). SHI score (continuous) was the only significant independent predictor of sleep quality on logistic regression (OR = 1.13 per unit increase; 95% CI: 1.08–1.19; p < 0.001), equivalent to a 3.4-fold increase in odds per 10-unit rise in SHI score. Female sex was additionally identified as a significant predictor (OR = 1.88; 95% CI: 1.00–3.53; p = 0.049). A significant positive correlation was observed between PSQI and SHI scores (r = 0.359, p < 0.001).

**Conclusion:** Poor sleep hygiene is highly prevalent among UMST medical students and is the most significant modifiable predictor of poor sleep quality, with each unit increase in SHI score increasing the odds of poor sleep quality by 13%. These findings highlight a gap in sleep health education within Sudanese medical institutions and support the integration of targeted sleep hygiene interventions into the medical curriculum.

## 1. INTRODUCTION

Sleep is a fundamental physiological process essential for cognitive function, immune regulation, metabolic homeostasis, and emotional wellbeing. For medical students — who face uniquely demanding academic and clinical schedules — adequate sleep is critical not only for academic performance but for the development of safe clinical reasoning. Evidence consistently demonstrates that sleep deprivation impairs cognitive function and increases the risk of medical errors, with downstream consequences for patient safety [1].

Sleep quality is substantially influenced by sleep hygiene: a set of behavioural and environmental practices that facilitate the initiation and maintenance of sleep. These include maintaining regular sleep and wake times, avoiding stimulants before bedtime, limiting use of the bed to sleep-related activities, and creating an optimal sleep environment [1]. While sleep hygiene awareness is necessary, adherence to these practices is what determines sleep outcomes.

Studies across medical schools in Saudi Arabia, Qatar, Ethiopia, Jordan, and Nigeria have reported poor sleep quality prevalence rates ranging from 32.5% to 80.3% among medical students, with poor sleep hygiene consistently emerging as a significant predictor [2–8]. However, no published study has examined sleep quality or sleep hygiene practices among medical students in Sudan. This gap is significant given Sudan’s distinct socioeconomic context, academic culture, and health system pressures.

This study aimed to address that gap by assessing sleep hygiene practices and their association with sleep quality among medical students at the University of Medical Sciences and Technology (UMST), Khartoum, Sudan — the first study of its kind in the country.

## 2. METHODS

### 2.1 Study Design and Setting

A facility-based cross-sectional study was conducted at the University of Medical Sciences and Technology (UMST), Khartoum, Sudan, a private medical university established in 1995. Data collection was carried out during the 2023–2024 academic year.

### 2.2 Study Population and Sampling

The target population comprised medical students in their 3rd, 4th, and 5th years of study (Batches 25, 26, and 27). Using Yamane’s formula with a margin of error of 0.05 and a population of 899 students, a sample size of 240 was calculated. Participants were selected using stratified random sampling, with each academic year as a stratum. Weighting by batch size yielded: 5th year (36.3%), 3rd year (35.4%), and 4th year (28.3%). Non-medical students and those unwilling to participate were excluded.

### 2.3 Data Collection Instruments

Two validated self-administered tools were used:

- Pittsburgh Sleep Quality Index (PSQI): A 19-item instrument producing a global score of 0–21. A score >5 indicates poor sleep quality (sensitivity 89.6%, specificity 86.5%).
- Sleep Hygiene Index (SHI): A 13-item instrument scored 0–52. A score >16 indicates poor sleep hygiene (sensitivity 77%, specificity 47.5%).

Both tools are validated and widely used in comparable student populations internationally. A sociodemographic section captured age, gender, year of study, marital status, and use of sleep aids.

### 2.4 Ethical Considerations

Ethical approval was obtained from the Scientific Research Committee of the Faculty of Medicine, UMST (approved 23 January 2024). Administrative permission was granted by the Dean of Student Affairs. All participants provided written informed consent, and data were anonymised and used for research purposes only.

### 2.5 Statistical Analysis

Data were entered in Microsoft Excel and analysed using SPSS version 23. Descriptive statistics were used for demographic and outcome variables. Independent sample t-tests and one-way ANOVA compared mean PSQI and SHI scores across demographic subgroups. Chi-square tests assessed associations between categorical variables and sleep quality outcome. Pearson correlation assessed the linear relationship between PSQI and SHI scores. Binary logistic regression identified independent predictors of poor sleep quality, with SHI entered as a continuous variable. Marital status was excluded from the regression model due to near-complete separation arising from the predominantly single study population, which precluded stable estimation. Statistical significance was set at p < 0.05.

## 3. RESULTS

### 3.1 Sociodemographic Characteristics

A total of 240 students participated (response rate 100%). The mean age was 21.7 years. Males constituted 51.7% (n=124) and females 48.3% (n=116). By year of study: 5th year 36.3% (n=87), 4th year 28.3% (n=68), 3rd year 35.4% (n=85). The majority (70%) reported no use of sleep aids; 15% used over-the-counter medications, 13.3% dietary supplements, and 1.7% prescription medication.

### 3.2 Sleep Quality

Poor sleep quality (PSQI >5) was found in 72.1% of participants, with a mean global PSQI score of 7.25 ± 2.66. Fifth-year students had the highest prevalence of poor sleep quality (79.3%), followed by 3rd year (72.9%) and 4th year students (61.7%). Female students had higher mean PSQI scores (7.6 ± 2.6) compared to males (6.9 ± 2.6).

Common sleep disturbances included short sleep duration (<6 hours) in 37.9% of students and sleep onset latency >30 minutes in 32.1%. Despite these findings, 66.3% of students subjectively rated their sleep as good, suggesting limited awareness of sleep quality deficits.

### 3.3 Sleep Hygiene

Poor sleep hygiene (SHI >16) was reported by 92.5% of participants (222/240), with a mean SHI score of 27.1 ± 7.9. The most frequently reported poor sleep hygiene practices were: engaging in stimulating activities before bedtime (66.6%), irregular bedtimes (61%), using the bed for non-sleep activities such as studying or watching television (60.9%), and planning or worrying in bed (60%). Table 2 presents the full frequency distribution of individual SHI items.

**Table 2.** Frequency distribution of Sleep Hygiene Index (SHI) items among UMST medical students (N=240).

| Sleep Hygiene Practice | Never<br>n(%) | Rarely<br>n(%) | Sometimes<br>n(%) | Frequently<br>n(%) | Always<br>n(%) |
| --- | --- | --- | --- | --- | --- |
| 1. Daytime naps $\geq 2$ hours | 27<br>(11.3) | 51 (21.3) | 76 (31.7) | 50 (20.8) | 36 (15.0) |
| 2. Irregular bedtimes | 2 (0.8) | 26 (10.8) | 63 (26.3) | 90 (37.5) | 59 (24.6) |
| 3. Irregular wake times | 8 (3.3) | 38 (15.8) | 68 (28.3) | 77 (32.1) | 49 (20.4) |
| 4. Exercise within 1hr of bedtime | 131<br>(54.6) | 50 (20.8) | 30 (12.5) | 22 (9.2) | 7 (2.9) |
| 5. Staying in bed longer than needed | 19 (7.9) | 31 (12.9) | 63 (26.3) | 74 (30.8) | 53 (22.1) |
| 6. Tobacco/caffeine within 4hrs of bedtime | 97<br>(40.4) | 29 (12.1) | 36 (15.0) | 43 (17.9) | 35 (14.6) |
| 7. Stimulating activities before bedtime | 14 (5.8) | 20 (8.3) | 46 (19.2) | 69 (28.7) | 91 (37.9) |
| 8. Going to bed stressed/angry/upset | 24<br>(10.0) | 45 (18.8) | 107 (44.6) | 49 (20.4) | 15 (6.3) |
| 9. Using bed for non-sleep activities | 13 (5.4) | 25 (10.4) | 56 (23.3) | 64 (26.7) | 82 (34.2) |
| 10. Uncomfortable bed | 68<br>(28.3) | 72 (30.0) | 61 (25.4) | 28 (11.7) | 11 (4.6) |
| 11. Uncomfortable bedroom | 68<br>(28.3) | 73 (30.4) | 56 (23.3) | 34 (14.2) | 9 (3.8) |
| 12. Important work before bedtime | 26<br>(10.8) | 44 (18.3) | 79 (32.9) | 73 (30.4) | 18 (7.5) |
| 13. Thinking/planning/worrying in bed | 18 (7.5) | 25 (10.4) | 53 (22.1) | 76 (31.7) | 68 (28.3) |

### 3.4 Associations Between Demographic Variables and Sleep Outcomes

Table 3 summarises the association between demographic variables and sleep quality and sleep hygiene scores.

**Table 3.** Association of demographic variables with PSQI and SHI scores. *Statistically significant (p < 0.05).

| Variable | Mean PSQI (p-value) | Mean SHI (p-value) | Sleep Quality Outcome ($\chi^2$, p) |
| --- | --- | --- | --- |
| Gender | p = 0.107 | p = 0.240 | p = 0.016* |
| Year of Study | p = 0.049* | p = 0.630 | p = 0.053 |
| Age | p = 0.012* | p = 0.040* | p = 0.170 |
| Marital Status | p = 0.040* | p = 0.645 | p = 0.209 |
| Type of Sleep Aid | p = 0.013* | p = 0.018* | p = 0.066 |
| SHI Group | — | — | p < 0.001* |

### 3.4 Correlation Between Sleep Quality and Sleep Hygiene

Pearson correlation analysis demonstrated a significant positive correlation between PSQI and SHI scores (r = 0.359, p < 0.001), indicating that students with poorer sleep hygiene practices had significantly poorer sleep quality.

### 3.6 Predictors of Sleep Quality

Binary logistic regression was performed with poor sleep quality as the dependent variable and age, gender, year of study, type of sleep aid used, and SHI score (continuous) as independent variables. Marital status was excluded due to near-complete separation in this predominantly single sample. The overall model accuracy was 74.6%, with a Nagelkerke R^2^ of 0.228.

SHI score was the only statistically significant independent predictor of poor sleep quality (β = 0.123, OR = 1.13, 95% CI: 1.08–1.19, p < 0.001). Each one-unit increase in SHI score increased the odds of poor sleep quality by 13%, equivalent to a 3.4-fold increase in odds per 10-unit rise (OR = 3.41, 95% CI: 2.09–5.57). Female sex was additionally a significant predictor (β = 0.631, OR = 1.88, 95% CI: 1.00–3.53, p = 0.049). Age, year of study, and type of sleep aid used were not significant predictors.

## 4. DISCUSSION

This study is, to our knowledge, the first to examine sleep quality and sleep hygiene practices among medical students in Sudan. The prevalence of poor sleep quality (72.1%) observed at UMST is consistent with rates reported in comparable studies from Saudi Arabia (76.4%), Qatar (70%), and Tunisia (72.5%), and substantially higher than those from Ethiopia (48.1%), Pakistan (65.4%), and India (49.2%) [2–8]. The variability across settings likely reflects differences in academic culture, socioeconomic pressures, study design, and cultural attitudes toward sleep.

The prevalence of poor sleep hygiene (92.5%) is among the highest reported in the literature for medical student populations. The most common problematic behaviours — stimulating activities before bedtime, irregular sleep schedules, and using the bed for non-sleep activities — are consistent with findings from Qatar, Saudi Arabia, and Ethiopia [3,6,8], suggesting these represent cross-cultural challenges in the medical student context.

Sleep hygiene emerged as the only significant independent predictor of sleep quality on logistic regression (OR = 1.13 per unit increase in SHI score, p < 0.001), with each 10-unit increase in SHI score associated with a 3.4-fold increase in the odds of poor sleep quality (OR = 3.41, 95% CI: 2.09–5.57). This magnitude of effect is consistent with findings from India (OR = 4.2) and Qatar (OR = 4.0) [5,6], reinforcing the central role of sleep hygiene as a modifiable target for intervention. Female sex was additionally a significant predictor (OR = 1.88, p = 0.049), consistent with prior evidence of gender differences in sleep quality [3,6].

The finding that fifth-year students had the worst sleep quality (mean PSQI 7.76) is consistent with international literature and likely reflects increased academic pressure associated with final MBBS examinations [7,8]. The gender difference — with females reporting higher mean PSQI scores (7.6 vs 6.9) — is similarly consistent with prior evidence and may relate to differential stress responses, social expectations, and study habits [3,6].

A notable discrepancy was observed between objective and subjective sleep quality: 66.3% of students subjectively rated their sleep as good despite 72.1% scoring above the PSQI threshold for poor sleep quality. This pattern — documented in Qatar and elsewhere — suggests limited awareness of what constitutes healthy sleep and highlights the need for structured sleep education within medical curricula [6].

### Limitations

This study is limited by its cross-sectional design, which precludes causal inference. The sample was drawn from a single institution, limiting generalisability to other Sudanese medical schools. Student knowledge and attitudes toward sleep hygiene were not assessed, and data were entirely self-reported. Marital status could not be included in the regression model as a covariate due to near-complete separation (>95% of participants were single), which precluded stable estimation; future studies with more heterogeneous populations should include this variable. Future research should employ multi-site designs, include objective sleep measures (e.g., actigraphy), and assess the impact of educational interventions on sleep hygiene adherence.

## 5. CONCLUSION

Poor sleep quality and poor sleep hygiene are highly prevalent among UMST medical students, with SHI score representing the single most significant modifiable predictor of sleep quality outcomes — each 10-unit increase in SHI score associated with a 3.4-fold increase in the odds of poor sleep quality. Female sex was additionally identified as a significant predictor. These findings establish a baseline for sleep health research among Sudanese medical students and provide evidence to support the integration of sleep hygiene education into the medical curriculum. Gender-specific interventions targeting female students are particularly warranted.

## DECLARATIONS

### Ethical Approval

Ethical approval was granted by the Scientific Research Committee, Faculty of Medicine, UMST (23 January 2024). Administrative approval was obtained from the Dean of Student Affairs, UMST.

### Informed Consent

Written informed consent was obtained from all participants prior to data collection.

### Competing Interests

The author declares no competing interests.

### Funding

No external funding was received for this study.

### Data Availability

The data supporting the findings of this study are available from the corresponding author upon reasonable request.

## Notes

### Competing Interest Statement

The authors have declared no competing interest.

### Funding Statement

This study did not receive any funding

### Author Declarations

Ethical approval was obtained from the Scientific Research Committee of the Faculty of Medicine, University of Medical Sciences and Technology (UMST) (approved 23 January 2024). Administrative permission was granted by the Dean of Student Affairs. All participants provided written informed consent, and data were anonymised and used for research purposes only.

## REFERENCES

1. Al-Kandari S, et al. Association between sleep hygiene awareness and practice with sleep quality among Kuwait University students. Sleep Health. 2017;3(5):342–7.

2. Alshahrani M, Al Turki Y. Sleep hygiene awareness: its relation to sleep quality among medical students at King Saud University. J Fam Med Prim Care. 2019;8(8):2628–32.

3. Molla A, Wondie T. Magnitude of poor sleep hygiene practice and associated factors among medical students in Ethiopia. Sleep Disord. 2021;2021:6611338.

4. DŽaferović A, Ulen K. Sleep habits among medical students and correlation between sleep quality and academic performance. Eur J Public Health. 2018;28(suppl_4).

5. Begum M, Puchakayala DSC. Study to determine prevalence of poor sleep quality and its correlation with sleep hygiene practices among medical students. Asian J Med Sci. 2022;13(9):151–5.

6. Ali RM, Zolezzi M, Awaisu A, Eltorki Y. Sleep quality and sleep hygiene behaviours among university students in Qatar. Int J Gen Med. 2023;16:2427–39.

7. Eze C. Sleep health among medical students in Abakaliki Nigeria: a descriptive study. Sleep Med X. 2024;7:100103.

8. Safhi MA, et al. The association of stress with sleep quality among medical students at King Abdulaziz University. J Fam Med Prim Care. 2020;9(3):1662–7.

9. Alwhaibi M, Al Aloola NA. Associations between stress, anxiety, depression and sleep quality among healthcare students. J Clin Med. 2023;12(13).

10. Nsengimana A, et al. Sleep quality among undergraduate medical students in Rwanda: a comparative study. Sci Rep. 2023;13(1):265.

